# Prevalence and characteristics of hazardous and harmful drinkers receiving general practitioners’ brief advice on and support with alcohol consumption in Germany: results of a population survey

**DOI:** 10.1101/2022.04.25.22274258

**Authors:** Sabrina Kastaun, Claire Garnett, Stefan Wilm, Daniel Kotz

**Affiliations:** Institute of General Practice, Addiction Research and Clinical Epidemiology Unit, Centre for Health and Society, Medical Faculty of Heinrich-Heine-University Düsseldorf, Germany; Institute of General Practice, Patient-Physician Communication Research Unit, Centre for Health and Society, Medical Faculty of Heinrich-Heine-University Düsseldorf, Germany; Department of Behavioural Science and Health, University College London, London, UK; SPECTRUM Consortium, London, UK

**Keywords:** general practitioner, primary care, brief advice, hazardous drinking, alcohol

## Abstract

**Objective:** The German treatment guideline on alcohol-related disorders recommends that general practitioners (GPs) offer brief advice on, and support with, reducing alcohol consumption to hazardous (at risk for health events) and harmful (exhibit health events) drinking patients. We aimed to estimate the implementation of this recommendation using data from the general population in Germany.

**Design:** Cross-sectional analysis of data (2021/2022) of a nationally representative household survey.

**Setting:** Population of Germany.

**Participants:** Representative sample of 2,247 adult respondents (>18 years) who reported hazardous or harmful drinking according to the Alcohol Use Disorders Identification Test-Consumption (AUDIT-C; score females: 4-12, males: 5-12).

**Main outcome measure:** Ever receipt of “brief GP advice on, or support with, reducing alcohol consumption”. Differences in the likelihood of ever receiving advice and/or support (yes/no) relative to respondents’ sociodemographic, smoking, and alcohol consumption characteristics were estimated using logistic regressions.

**Results:** Ever receipt of GP advice on/support with reducing alcohol was reported among 6.3% (95%CI=5.3%-7.4%), and the offer of support among 1.5% (95%CI=1.1%-2.1%) of the hazardous and harmful drinking respondents. The likelihood of having ever received advice/support was positively associated with being older (odds ratio (OR)=1.03 per year, 95%CI=1.01-1.04), a current or former (versus never) smoker (OR=2.36, 95%CI=1.46-3.80; OR=2.17, 95%CI=1.23-3.81), and with increasing alcohol consumption (OR=1.76 per score, 95%CI=1.59-1.95). One in two harmful drinking respondents (AUDIT-C score 10-12) reported appropriate advice/support. The likelihood was negatively associated with being female (e.g., OR=0.32, 95%CI=0.21-0.48), having a medium and high (versus low) education, and with increasing household income.

**Conclusions:** A small proportion of people drinking at hazardous and harmful levels in Germany report having ever received brief GP advice on, or support with, reducing alcohol consumption. The implementation of appropriate advice or support seems to be strongly linked to specific sociodemographic characteristics, tobacco smoking, and the alcohol consumption level.

**Strengths and limitations of this study:**

- The principal strength of this study is the large, nationally representative population sample.
- The cross-sectional study design and temporality issues with our measures (alcohol consumption was measured with reference to the present and the outcome as “ever receipt of GP advice or support”) limited our ability to explore causal relationships.
- The outcome measure had a complex, not entirely hierarchical structure, which may have led to respondents being unsure of which response option to select.
- Data were collected during the COVID-19 pandemic, during which alcohol consumption in Germany seemed to have changed. It is unclear how this might have influenced GPs’ behaviour.
- Due to the socially loaded topic, respondents may not have answered truthfully or repressed a previous conversation with their GP on their alcohol consumption. We did not assess the GPs’ view on the topic.

## INTRODUCTION

Hazardous drinking is defined as a quantity or a repeated pattern of alcohol consumption that places a person at risk for adverse health events, whereas harmful drinking verifiably results in such events.[1-3] Alcohol dependence, on the other hand, is seen as a complex drinking pattern, characterised by persistent consumption despite harmful consequences, craving, the prioritisation of drinking over other activities, tolerance development, and withdrawal symptom.[1-3] Although this terminology is now commonly used and acknowledges the spectrum of risk that tends to increase with increasing drinking, universal cut-offs with regard to consumption levels and associated risks are missing, making a clear distinction of these theoretical drinking patterns often difficult in practice.[1-3]

Alcohol misuse contributes to around 3 million deaths each year globally, and is responsible for around 5.1% of global disability-adjusted life-years.[4] Germany ranks above the average level in the European Union of pure alcohol consumption per capita and year in the adult population (13.4 litres of pure alcohol versus 9.8 litres).[4] Latest nationally representative prevalence data show that around 20% of the adult population consume alcohol at least at a hazardous level,[5] including a smaller proportion (approximately 3% per group,[6]) of harmful and alcohol dependent drinkers. There is strong evidence of a dose-response relationship between alcohol consumption and alcohol-related harms,[7, 8] whereby any person drinking at a hazardous level or above would benefit from reducing their alcohol consumption.

The implementation of brief interventions in primary healthcare settings is both an effective [9-13] and cost-effective [4] approach to reducing hazardous and harmful drinking. Brief interventions usually include feedback on consumption, brief advice to reduce or quit drinking, motivational enhancement and goal setting, and further support such as referral to specialised treatment or the development of a personal reduction plan.[10, 14]

General practitioners (GPs) are well placed to address alcohol use disorders as they commonly see patients of various ages with a broad range of (alcohol-related) health conditions,[14] and the long-lasting patient-GP relationship can help to reduce feelings of stigmatisation and irritation in the patient.[2] The current German clinical guideline on the treatment of alcohol-related disorders [2] recommends that brief interventions should be offered to hazardous and harmful drinkers in the primary care setting. Identification of patients can be carried out by means of screening and pragmatic case-finding (i.e., when the issue is raised by the patient or when the GP notices alcohol-related conspicuousness).[2] The latter seems to be common in the GP setting.[15]

For individuals with alcohol dependence, evidence on the effectiveness of brief interventions is inconsistent, but according to the German treatment guideline, brief interventions can be a sensible measure among this group for ethical and pragmatic reasons, if other and more intensive interventions (e.g., specialised inpatient treatment) are rejected by the patient.[2]

Addressing a patient’s alcohol consumption is still perceived as an emotionally difficult and socially loaded issue. Despite strong evidence on the effectiveness of brief interventions, international data suggest that its implementation in routine GP practice remains challenging.[16-19] A population survey from England comparing GP advice on drinking with smoking found that only 6.5% of hazardous and harmful drinkers received advice on their alcohol consumption during the past year, compared with half of smokers that received advice on quitting smoking.[18] Although most primary care clinicians report that they ask their patients about alcohol use, far fewer offer advice or recommend treatment.[20] A survey which used routine GP data showed that GPs struggled more often at identifying hazardous than dependent drinkers.[19] No representative, national data are available on the implementation of the German clinical guideline on hazardous and harmful drinking in GP settings. Only one study provides initial figures from a single federal state, Bremen, collected in 2016, where 2.9% of all hazardous drinking patients were screened by their GPs, and 1.4% received a brief intervention.[21]

Up-to-date and representative national figures on the provision of brief alcohol interventions in GP settings are needed to be able to precisely inform health policy and – if needed – the development of interventions aiming at improving the implementation of treatment recommendations on brief alcohol interventions in primary care. It is also important to report whether the provision of brief alcohol interventions differs by recipients’ characteristics in order to identify potentially underserved groups of society. This study therefore aimed to explore the following questions using data from a nationally representative sample of adults (aged >18) in Germany who self-report hazardous or harmful drinking (operationalised using the Alcohol Use Disorders Identification Test-Consumption (AUDIT-C)).[22]

## RESEARCH QUESTIONS

Among adults in the population of Germany drinking at hazardous and harmful levels:

1. What proportion reports that a GP ever asked about their alcohol consumption, advised them to drink less, offered help or support with drinking less, and offered help with making use of external medical or psychological support because of alcohol consumption?
2. What proportion report ever (versus never) receiving brief GP advice on, or support with, reducing alcohol consumption stratified by person characteristics: age, sex, education, income, smoking status, migration background, region of residence, and the overall alcohol consumption?
3. Are there any differences in the likelihood of ever (versus never) receiving brief GP advice on, or support with, reducing alcohol consumption within each measured recipient characteristic?

Since the clinical guideline does not provide a clear recommendation on the exact level of alcohol consumption at which a brief intervention should take place,[2] our study will address individuals consuming alcohol at least at a hazardous level. From a preventive medicine perspective, we most likely expect an intervention from GPs among this group of risk.

## MATERIALS AND METHODS

### Study design

We used data from the cross-sectional German Study on Tobacco Use (DEBRA: “Deutsche Befragung zum Rauchverhalten”): an ongoing representative household survey on tobacco and nicotine product use in Germany (www.debra-study.info).[23] The study is conducted by a market research institute, has been registered at the German Clinical Trials Register (DRKS00011322, DRKS00017157), and received approval by the ethics committee of Heinrich-Heine-University Düsseldorf (HHU 5386R). Since 2016, the DEBRA study collects data every other month from computer-assisted, face-to-face household interviews of people aged >14.

### Study population

Data were aggregated from seven survey waves (waves 28-34) collected between February 2021 and February 2022 (N=14,327). Since January 2020, respondents are selected by using a dual frame design: a composition of random stratified sampling (50% of the sample) and quota sampling (50% of the sample). This sampling design has been described in detail elsewhere: osf.io/s2wxc/. Details on the general sample selection have been published in a study protocol.[23]

Alcohol consumption was measured with the AUDIT-C;[22] a three-item measure including questions on 1) frequency, 2) quantity, and 3) frequency of occasional heavy drinking; full details are given in **Table 1**. The AUDIT-C overall score indicates the level of alcohol consumption and ranges from 0 to 12.[22] As recommended in the German treatment guideline on alcohol-related disorders [2] and in underlying studies,[24, 25] a gender-specific AUDIT-C cut-off score of >5 in males, and of >4 in females was used to operationalise “at least hazardous drinking”. Respondents who answered that they never drink alcohol did not receive questions 2 and 3, and were excluded from the analysis. The study population included all adults (aged >18 years, n=14,026) who reported at least hazardous drinking, resulting in a total sample of n=2,712 (19.3%) hazardous or harmful drinkers. Respondents aged 14 to 17 were excluded from the analysis as 16 is the national legal age of sale for beverages containing <15% of alcohol by volume (ABV), and 18 is the legal age of sale for beverages with >15% ABV.

**TABLE 1.** Measures of assessing alcohol consumption and ever receiving brief GP advice on/support with reducing alcohol consumption

| Exposure measure: hazardous and harmful alcohol consumption (AUDIT-C items [22]) | Outcome measure:<br>Brief GP advice on alcohol consumption |
| --- | --- |
| 1. *How often do you have a drink containing alcohol?*<br>- never (score 0)<br>- at least once a month or less (score 1)<br>- 2–4 times per month (score 2)<br>- 2–3 times per week (score 3)<br>- 4+ times per week (score 4)<br>- no answer<br>2. *How many units of alcohol do you drink on a typical day when you are drinking?*<br>- 1–2 (score 0)<br>- 3–4 (score 1)<br>- 5–6 (score 2)<br>- 7–9 (score 3)<br>- 10+ (score 4)<br>- no answer<br>3. *How often have you had 6 or more units if female, or 8 or more if male, on a single occasion in the last year?*<br>- never (score 0)<br>- less than monthly (score 1)<br>- monthly (score 2)<br>- weekly (score 3)<br>- daily or almost daily (score 4)<br>- no answer | *Has a GP ever asked you about your alcohol consumption and, if so, recommended something to you in this regard?*<br>1. No, a GP has never asked me about my alcohol consumption<br>2. Yes, a GP has asked me about my alcohol consumption at some point<br>3. Yes, a GP has advised me to drink less at some point<br>4. Yes, a GP has offered me help or support to drink less at some point<br>5. Yes, a GP has advised or helped me to make use of psychological or medical support because of my alcohol consumption at some point<br>6. I don't remember if a GP has ever addressed my alcohol consumption<br>7. No answer |

### Outcome measure

Respondents were asked about “ever receipt of GP advice on, or support with, reducing alcohol consumption”, using a question that was adopted from previous studies on GP advice on smoking cessation in the German,[26] and in the Dutch and English population,[27] and which was critically reviewed by an experienced GP (see **Table 1**).

For further analyses, the response options for this question were dichotomised into ‘no’ (options 1-2; 0) and ‘yes’ (options 3-5; 1), see **Table 1**. Respondents who answered “I don’t remember” (n=443) or refused to answer (n=22) were excluded from the analyses.

### Exposure variables

The following sociodemographic characteristics were measured: age; sex (female versus male); region of residence (rural versus urban setting), migration background (yes versus no), current tobacco smoking status (current, former, never), and alcohol consumption as a continuous variable (AUDIT-C score with a possible range of 4-12 among women and of 5-12 among men who drink at least hazardously).

Alcohol consumption varies between regions of residence, with a majority of studies reporting higher drinking rates in rural communities.[28] We assumed that this somehow affects the awareness and behaviour of GPs with regard to advice on drinking. This variable was assessed by using the national classification of regions (“BIK Regionsklassifizierung”,[29]), consisting of five categories (e.g., metropolitan area or subcentres) which were dichotomised for the analyses: urban versus rural setting.

German population surveys suggest that individuals with migration background relative to those without consume less alcohol.[30] Respondents were asked: “Was one of your parents born abroad?”. Migration background applied if at least one parent did not have German nationality by birth.

Measured socioeconomic status (SES) variables were: educational qualification (low [9 years of education, or no graduation], medium [>10 years], high [>12 years]), and monthly net household income calculated per person (details on the calculation can be found here: https://osf.io/387fg/). Income was entered as a continuous variable coded from 0 (€0 income/month) to 7 (>€7,000/month) in the regression models. For descripitive purposes, income was categorised into: low (<20th percentile), medium (20th to 80th percentiles), and high (>80th percentile), approximately reflecting the distribution in the German population.

### Statistical analyses

The study protocol and analysis plan was written prior to analysing data and pre-registered on the Open Science Framework: osf.io/3fe87.

Data were analysed and reported unweighted as information on true population parameters of the population of hazardous drinkers were not available. Analyses were performed with IBM SPSS Statistics for Windows, Version 27.0 (Armonk, NY: IBM Corp).

To address **research question 1**, we report descriptive prevalence data on the various levels of GP advice on, or support with, reducing alcohol consumption as percentages together with 95% confidence intervals (95%CI).

To address **research question 2**, we present prevalence data including 95%CI of the dichotomous outcome variable “ever receipt of GP advice on, or support with, reducing alcohol consumption (=yes versus no)” stratified by all categorical exposure variables. For the continuous scaled AUDIT-C score we present a figure showing the prevalence of “ever receipt of GP advice/support (=yes)” in relation to the range of possible AUDIT-C scores among the sample of hazardous and harmful drinkers.

To address **research question 3**, a series of univariate logistic regression models were conducted to explore potential differences in the likelihood of ever (yes vs. no) receiving brief GP advice on, or support with, reducing alcohol consumption for all exposure variables. Regression models were adjusted for survey wave as a potential confounding variable.

A considerable number (16.3%) of respondents did not remember if a GP had ever addressed their alcohol consumption and some refused to answer (0.8%). For the analyses of research questions 2 and 3, this group was excluded. We compared whether this group differed systematically from respondents who provided an answer on our outcome question by using the chi-square test and Mann-Whitney U test (**Supplemental Table 1**).

#### Dealing with missing data

Missing data were sparse (<0.6% for all variables except for income (2.9%) and migration background (4.8%)), and data were analysed using complete cases. Due to an incorrect questionnaire instruction in four survey waves (28-31), 22.6% of respondents who reported to the first AUDIT-C question with “at least once a month or less” did not receive the AUDIT-C questions 2 and 3, and were not interviewed on the primary outcome. However, an analysis of AUDIT-C data from earlier waves of the DEBRA study [5] with the correct questionnaire instruction showed that only 1% of respondents who provided the same answer, were identified as hazardous or harmful drinkers. We thus assume that around 1% of hazardous or harmful drinkers were lost across four of the seven survey waves due to this mistake. These missing data were assumed to be completely at random, and thus excluded from the analyses.

## RESULTS

The final analytic sample consisted of all adult respondents who reported at least hazardous drinking, and provided an answer on, or could remember, whether or not a GP had advised them on reducing alcohol consumption (n=2,247). The sample is described in **Table 2**. The mean age of this group was 49.5 years (standard deviation (SD)=17.0), and 45.0% (n=1012) of the respondents were female. The mean AUDIT-C score was 5.9 (median: 6, standard deviation (SD)=1.3) for males and 4.8 (median: 4, SD=1.1) for females.

**TABLE 2.**
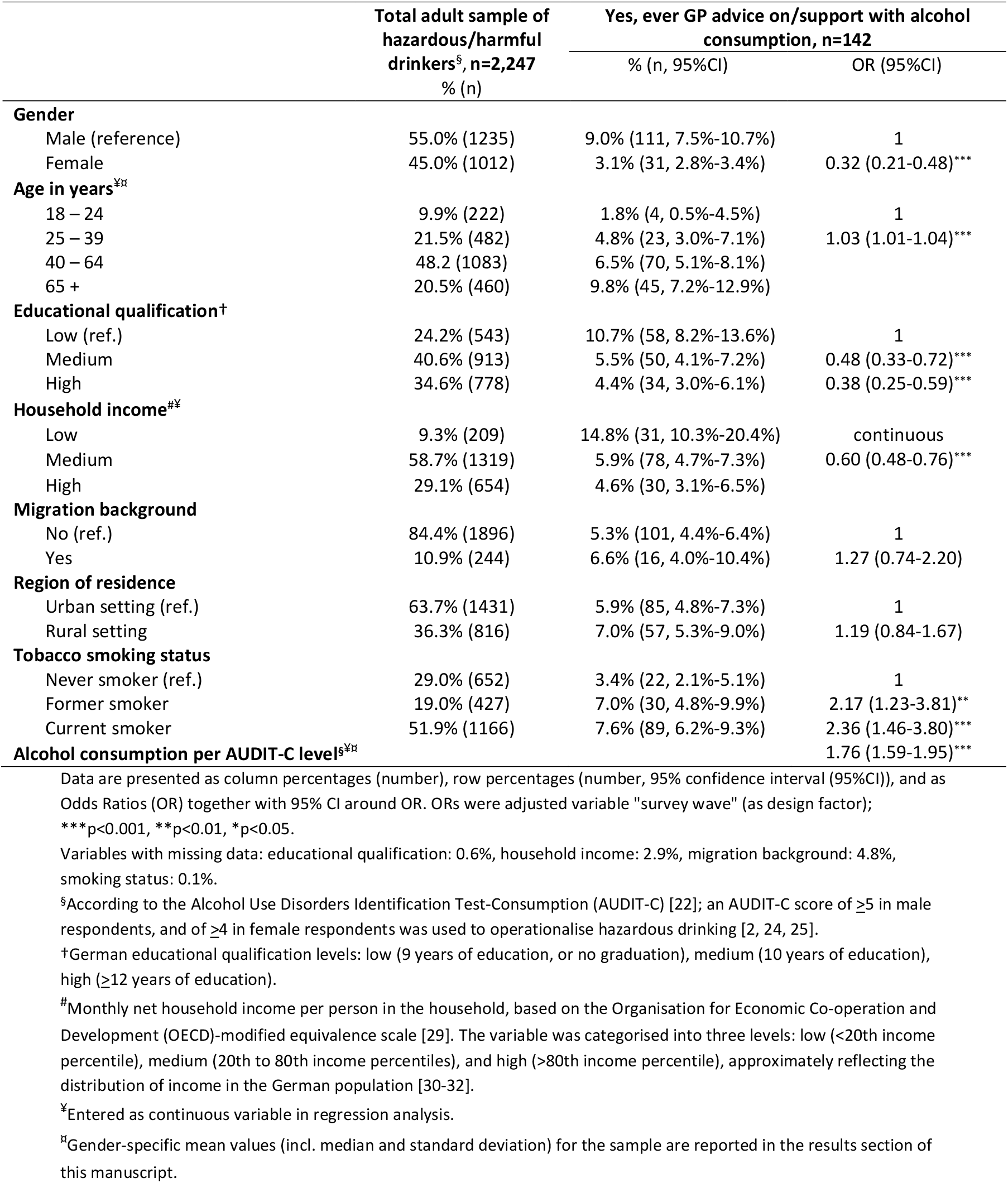
Characteristics of all adult hazardous or harmful drinking respondents (n=2,247), and prevalence estimates on the ever receipt of brief GP advice on/support with reducing alcohol consumption (=yes) relative to the respondents’ characteristics; including results of univariate regressions models on associations between these characteristics and ever receipt of GP advice.

Respondents who didn’t remember whether or not they received GP advice seem to be more often current smokers and lower educated but did not systematically differ from respondents who did remember (**Supplemental Table 1**).

### Proportion of hazardous or harmful drinking adults reporting various levels of GP advice/support

Among hazardous or harmful drinkers, 82.2% (95%CI=80.5%-83.7%; n=1,846) reported that a GP had never asked them about their alcohol consumption, 11.5% (95%CI=10.2%-12.9%; n=259) reported that they had ever been asked about drinking by a GP, and 6.3% (95%CI=5.3%-7.4%; n=142) said that a GP had ever advised them to drink less, or offered either help or support or advised or helped them to make use of medical or psychological support to drink less (see **Figure 1**). Such support was reported by 1.5% (95%CI=1.1%-2.1%) of the hazardous and harmful drinking respondents.

**Figure 1:**
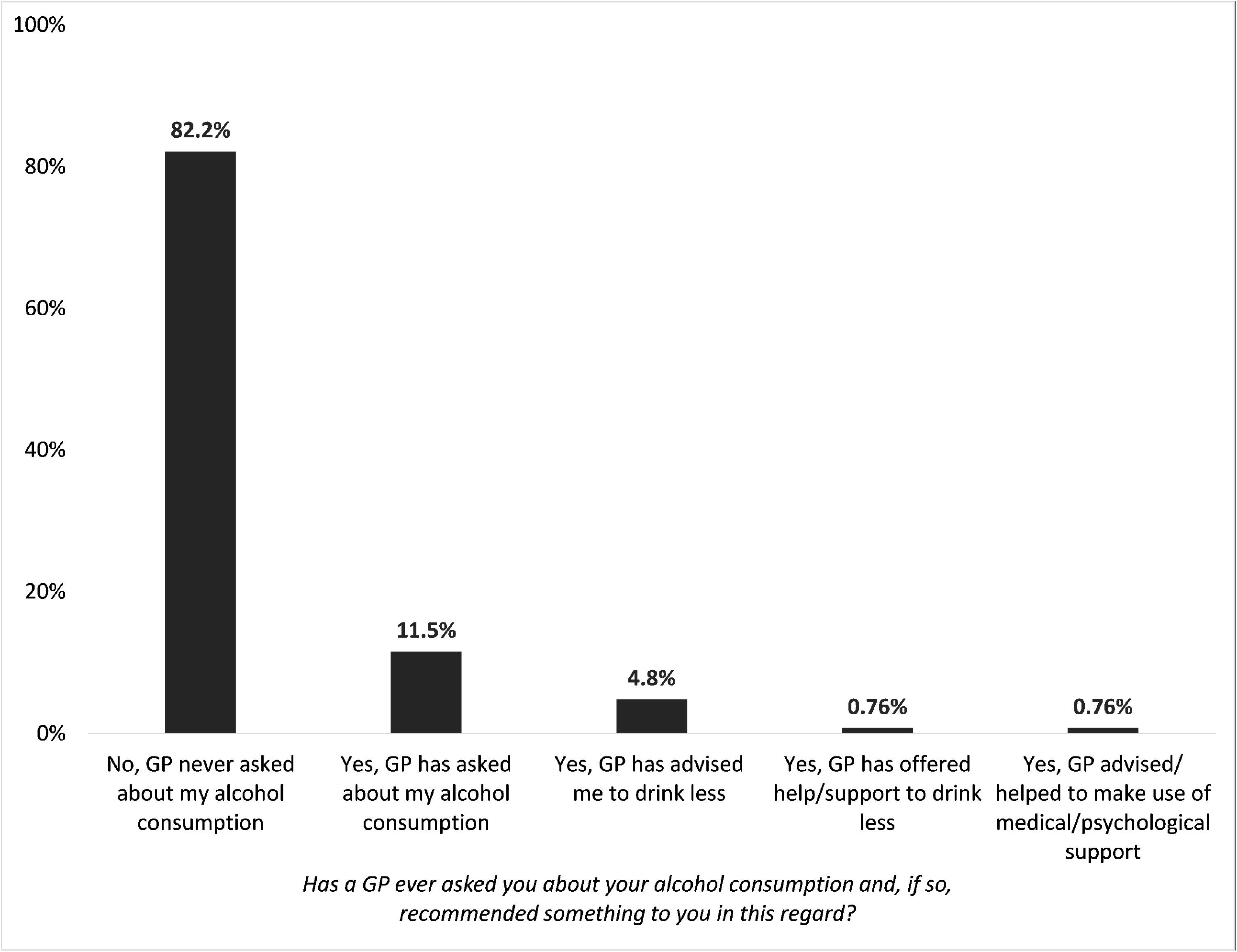
Prevalence estimates on the various levels of GP advice on, or support with, reducing alcohol consumption (self-reported) among the total sample of adult hazardous or harmful drinkers (n=2,247) reported as percentages together with 95% confidence intervals.

### Proportion of hazardous or harmful drinking adults reporting ever receipt of GP advice/support stratified by recipients’ characteristics

Men (compared with women), respondents of higher age (65+ years), those with low (compared with medium and high) education and household income, and current and former smokers (compared with never smokers) reported relatively more often to have ever received brief GP advice on, or support with alcohol consumption, see **Table 2**.

The higher the overall AUDIT-C score, the higher the rate of reporting ever receipt of GP advice on, or support with, reducing alcohol consumption, see **Figure 2**. In persons with an AUDIT-C score >9 – which is viewed as harmful or potentially dependent drinking pattern [37] – 51.5% (n=17/33) have ever received GP advice on, or support with reducing alcohol consumption.

**Figure 2:**
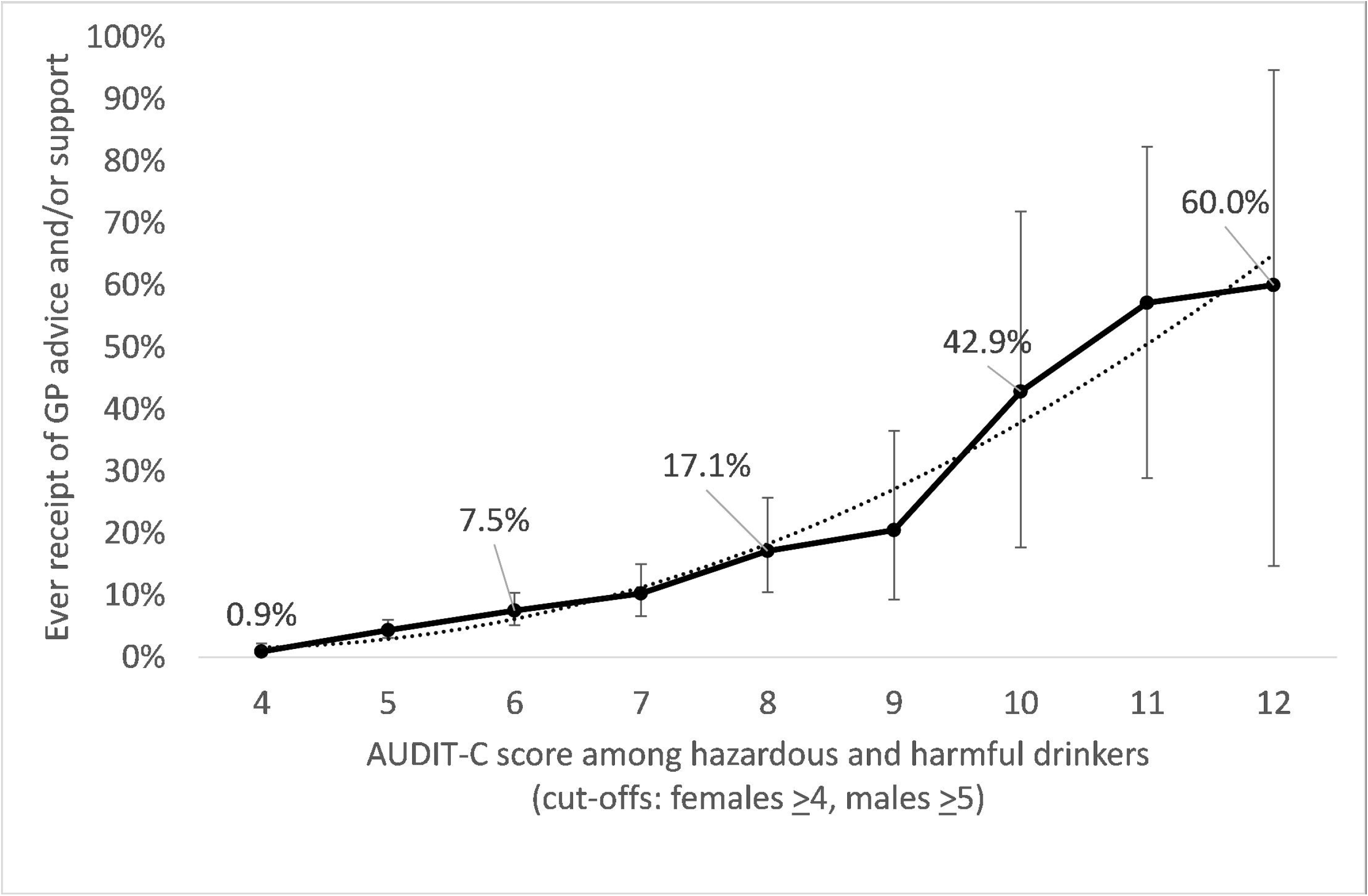
Ever receipt of GP advice on, or support with, reducing alcohol consumption (yes, self-reported) relative to the total AUDIT-C score among the total sample of adult hazardous or harmful drinkers (n=2,247) reported as percentages together with 95% confidence intervals (black line; dotted line: trend line, polynomial function, R^2^=0.97).

### Likelihood of ever receiving GP advice/support relative to recipients’ characteristics

The likelihood of ever receipt of brief GP advice on, or support with, reducing alcohol consumption was positively associated with being older, being a former or a current smoker, and reporting a higher alcohol consumption level at the time of the survey, see **Table 2**. The likelihood was negatively associated with being female, having medium and high (compared with low) educational qualification, and with increasing monthly household income.

No significant differences were detected relative to the respondents’ migration background or region of residence.

## DISCUSSION

In a large representative sample of the general population of adults in Germany who report hazardous or harmful drinking, about 12% reported having ever been asked by a GP about their alcohol consumption, and about 6% reported having ever received GP advice to drink less, including 1.5% who were also offered support to drink less or to make use of psychological or medical assistance for that purpose. However, in the subgroup of people reporting harmful or potentially dependent drinking (AUDIT-C score 10-12,[31]) around half had ever received brief GP advice on, or support with, reducing alcohol consumption.

To our knowledge, this is the first nationwide study to estimate the implementation of the clinical guideline recommendation on the provision of GPs’ brief advice on, or support with alcohol consumption in the population of Germany. Our findings are broadly consistent with a previous study (not methodologically comparable) in a single federal state in Germany,[21] and a population survey in England (with comparable methodology).[18] Both studies indicate insufficient implementation of brief alcohol interventions in primary care.

In the current study there were significant differences in the likelihood of having ever received GP advice or offer of support by personal characteristics. Hazardous and harmful drinkers of older age, current or former smokers, and those with higher alcohol consumption had higher odds of reporting ever receipt of GP advice on, or support with alcohol consumption, whereas females (versus males) and respondents with higher income or medium and high (versus low) educational qualification had substantially lower odds. Higher likelihood of receiving GP advice on alcohol consumption among older respondents and among current or former smokers might be associated with awareness among GPs of existing (alcohol-related) health conditions and of polysubstance use, as well as with demand for treatment among patients. The implementation of alcohol screening and brief intervention in individuals with co-morbidities seems to be largely accepted among GPs and higher than in those without co-morbidities.[32-35]

Our findings suggest that hazardous and harmful drinking women and those with higher SES are under-recognized by GPs when it comes to brief alcohol intervention. This is concerning as, although prevalence of hazardous and harmful drinking is higher among men, yet around one in 10 women report drinking at least at a hazardous level.[5] In addition, evidence is good that individuals with higher compared to with lower SES may consume similar or even greater amounts of alcohol and show higher prevalence rates of hazardous drinking.[5, 36] Gender gaps in GP-delivered alcohol interventions have been reported before [18, 37, 38] but we can only speculate on underlying reasons. This could be due to greater concerns about stigmatisation or shame, leading women and individuals with higher SES less often admit alcohol use to their GPs. On the other hand, implicit cognitive bias and stereotypes might influence the GPs’ decision on who to screen for alcohol misuse.[39] Another possible explanation might be that specific groups of society are less likely to consult a GP, which has been reported for higher SES groups,[40, 41] though is not the case for women who tend to visit their GP more often than men.[42]

### Implications for policy and practise

GPs are a major force to improve the prevention of alcohol-related harm on a population level, and as this study shows, they already intervene in about half of harmful drinking patients. However, from a preventive medicine perspective, this study reveals a need to improve the implementation of guideline recommendations for hazardous drinking in the GP setting. Whilst the clinical guideline does not provide a clear recommendation on the exact level of alcohol consumption at which a brief intervention should take place,[2] alcohol-related harms are dose-dependent and therefore it is important to provide brief interventions for all individuals at alcohol-related risk. Previous studies showed that education and post-graduate training predicts the GP delivery of brief alcohol interventions to hazardous drinkers,[32] and that training GPs can significantly increase alcohol screening and brief intervention rates in primary care.[43] This is particularly the case when being tailored to the barriers and facilitators towards the implementation of such interventions in the GP setting and when being developed on the basis of a behaviour change theory.[44] Further synergistic effects have been shown when financial incentives, training and support were offered together.[45]

In Germany, appropriate training is not offered by default, neither during medical education nor as post-graduate training for physicians. As a consequence, many GPs [46], as well as medical students,[47] in Germany do not feel adequately trained to diagnose and treat patients with alcohol problems. This lack of training had also been identified as a major barrier towards the routine implementation of brief alcohol intervention in primary care in the United Kingdom.[16]

### Strengths and limitations

A major strength of this study is the large, nationally representative sample. However, there are also limitations. First, data was self-reported, introducing risk for recall bias that may have affected the prevalence estimates, most likely resulting in an under estimate. Secondly, the cross-sectional study design and that GP advice and support was measured as “ever receipt”, whereas alcohol consumption was measured with approximate reference to the present, limited our ability to explore causal relationships. Comparable temporality issues might also occur for some of the sociodemographic characteristics such as income or place of residence. Based on this, we did not conduct multivariable regression analyses, and our results are not adjusted for potential confounding through interaction effects between the exposure variables. Future research should look to estimate the potential causal effect of someone’s characteristics and the likelihood of ever receipt of GP advice on, or support with, reducing alcohol consumption.

Thirdly, the question on “ever receipt of GP advice and support” had a complex structure (participants are asked about GP advice on as well as different types of support with drinking less), which could have led to difficulties in understanding. However, the face-to-face interview method and support available from the interviewers may have mitigated this risk. In addition, this measure did not follow an exact hierarchical structure and there was no clear distinction between “internal support” (e.g., offered by the GP), and “external support” (e.g. psychosocial services). This may have led to respondents being unsure of which response option to select. Fourthly, data were collected during the COVID-19 pandemic, during which alcohol consumption in Germany seemed to have decreased on average but increased in specific subgroups of the population.[48, 49] It is unclear how this might have influenced the behaviour of GPs. Finally, due to the socially loaded topic, respondents may not have answered truthfully or repressed a previous conversation with their GP on the topic. It is therefore important to also assess the GPs’ view on the topic, including in-depth information on barriers to the implementation of the treatment guideline recommendations in their daily practice. Previous surveys among GPs, however, usually only assessed how often, in general, GPs screen for alcohol or provide brief intervention, but not in relation to the number of their hazardous drinking patients.[32, 50]

## Conclusion

Our findings suggest that hazardous drinking – that places a person at risk for adverse health events – does not seem to be adequately addressed by GPs in Germany, particularly among women and individuals from higher SES groups. From a preventive medicine perspective, this results in missed opportunities to reduce alcohol-related harm. However, the probability of having ever received brief alcohol intervention by a GP increases with increasing drinking levels. Around every second harmful drinking person seems to have ever received brief GP advice on, or support with, reducing alcohol consumption. Although we did not analyse causal relationships, this study is a call for action in order to further explore underlying reasons why clinical guideline recommendations on brief alcohol interventions are not implemented more often in the German general practice setting, as well as to explore potential approaches for improvement.

## Data Availability

The data underlying this study are third-party data and are available to researchers on reasonable request from the corresponding author. All proposals requesting data access will need to specify how it is planned to use the data, and all proposals will need approval of the DEBRA study team before data release.

## Statements

## Acknowledgements

The authors thank Erika Baum for her constructive feedback on the outcome measure (the question on GP advice on alcohol consumption) as well as on the first version of the analysis protocol. The authors also thank Constanze Cholmakow-Bodechtel and Franziska Wenng from the market research institute “Kantar” for the collection of the data.

## Statement of Ethics

The study protocol has been peer-reviewed and approved by the ethics committee of Heinrich-Heine-University Duesseldorf, Germany (HHU 5386/R). The fieldwork is conducted by the market research institute Kantar, Germany. Interviewers from Kantar make sure that all participants give oral informed consent. This method of consent has been approved by the ethics committee.

## Patient and Public Involvement statement

Patients or the public were not involved in the design, or conduct, or reporting, or dissemination plans of this research.

## Author Contributions

SK coordinates the DEBRA study, conceptualised and drafted the analysis protocol, drafted the manuscript, analysed and interpreted the data. CG and SW: provided expert advice on the study protocol and critically revised the analysis protocol and the manuscript. DK conceived the DEBRA study, supervised the analyses, and critically revised the analysis protocol and the manuscript. All named authors contributed substantially to the manuscript and agreed on its final version.

