## Supplementary Table 1 for "Prevalence and characteristics of hazardous and harmful drinkers receiving general practitioners’ brief advice on and support with alcohol consumption in Germany: results of a population survey"

**SUPPLEMENTARY TABLE 1.** Characteristics of all adult hazardous or harmful drinking respondents who remembered whether they received GP advice or not (n=2,247) compared to those who didn’t remember or refused to answer (n=465)

|  | Respondents who remember  n=2,247, % (n) | | | Respondents who did not remember/refused to answer, n=465, % (n) | | *p*-value and  test statistics^a^ |
| --- | --- | --- | --- | --- | --- | --- |
| Gender |  | | |  | |  |
| Male | 55.0% (1235) | | | 52.5% (244) | | *Χ^2^*(1)=1.0, *p*=0.327 |
| Female | 45.0% (1012) | | | 47.5% (221) | |  |
| Age in years | |  |  | |  |  |
| 18 – 24 | 9.9% (222) | | | 8.0% (37) | | *Χ^2^*(3)=4.0, *p*=0.263 |
| 25 – 39 | 21.5% (482) | | | 20.0% (93) | |  |
| 40 – 64 | 48.2% (1083) | | | 48.2% (224) | |  |
| 65 + | 20.5% (460) | | | 23.9% (111) | |  |
| Educational qualification† | |  |  | |  |  |
| Low | 24.3% (543) | | | 27.4% (125) | | *Χ^2^*(2)=3.3, *p*=0.197 |
| Medium | 40.9% (913) | | | 41.8% (191) | |  |
| High | 34.8% (778) | | | 30.9% (141) | |  |
| Person net household income class^#^ | |  |  | |  |  |
| Low | 9.6% (209) | | | 12.5% (57) | | *Χ^2^*(2)=7.1, *p*=0.029* |
| Medium | 60.4% (1319) | | | 62.8% (287) | |  |
| High | 30.0% (654) | | | 24.7% (113) | |  |
| Migration background |  | | |  | | *Χ^2^*(1)=0.2, *p*=0.653 |
| No (ref.) | 88.6% (1896) | | | 89.3% (394) | |  |
| Yes | 11.4% (224) | | | 10.7% (47) | |  |
| Region of residence |  | | |  | | *Χ^2^*(1)=1.5, *p*=0.222 |
| Urban setting (ref.) | 63.7% (1431) | | | 66.7% (310) | |  |
| Rural setting | 36.3% (816) | | | 33.3% (155) | |  |
| Tobacco smoking status | |  |  | |  |  |
| Current smoker | 29.0% (652) | | | 36.4% (168) | | *Χ^2^*(2)=12.3, *p*=0.002** |
| Former smoker | 19.0% (427) | | | 20.0% (92) | |  |
| Never smoker | 51.9% (1166) | | | 43.6% (201) | |  |
| Alcohol consumption^§^, MD (IQR) | 5 (5-6) | | | 5 (5-6) | | z=-0.455, p=.649 |

Data are presented as column percentages (number), unless stated otherwise; MD = median, SD = standard deviation, **p<0.01, *p<0.05. Differences when calculating the total percentage can be explained by sparse missing data on the respective variable.

^a^Results of Chi-square (*Χ^2^*) test and Mann-Whitney U test (*z*).

^#^Monthly net household income per person in the household, based on the Organisation for Economic Co-operation and Development (OECD)-modified equivalence scale [29]. The variable was categorised into three levels: low (<20th income percentile), medium (20th to 80th income percentiles), and high (>80th income percentile), approximately reflecting the distribution of income in the German population [30-32].

†German educational qualification levels: low (9 years of education, or no graduation), medium (10 years of education), high (≥12 years of education).

^§^According to the Alcohol Use Disorders Identification Test-Consumption (AUDIT-C) [22]; an AUDIT-C score of ≥5 in male respondents, and of ≥4 in female respondents was used to operationalise hazardous drinking [2, 24, 25].
